# One in Five Reported Catheter-Associated Urinary Tract Infections Is Not Treated: An Opportunity to Align Surveillance With Antimicrobial Stewardship

**DOI:** 10.64898/2026.11.26.26368998

**Authors:** Valeria Fabre, Avinash Gadala, Larry Baker, Adina Ivanica, Erin Barker, Sara C. Keller, Lisa L. Maragakis, Sara E. Cosgrove

**Author notes:** **Corresponding author:** Valeria Fabre, MD, Division of Infectious Diseases, Johns Hopkins University School of Medicine, 600 North Wolfe Street, Halsted 8, Rm 840, Baltimore, Maryland 21287.

## Abstract

The National Healthcare Safety Network (NHSN) catheter-associated urinary tract infection (CAUTI) surveillance definition may not correlate well with clinical infection.

**Objective:** To describe the frequency of antibiotic treatment of NHSN-reported CAUTIs and identify factors associated with non-treatment.

**Methods:** Retrospective study of adults with NHSN-reported CAUTI at five hospitals (two teaching, three community) in Maryland and Washington, DC, January 2023–May 2026. Antibiotic indication and alternative diagnoses for symptoms were extracted from the electronic health record by chart review.

**Results:** Of 301 CAUTI events, 63 (20.9%) were not treated with antibiotics. Fever was the qualifying criterion for CAUTI in 244 (81.1%), localizing urinary findings were present in only 62 (20.60%); and the urinalysis lacked pyuria (WBC <10/HPF or ≤50/µL) for 37 patients (12.3%). There was a greater proportion of untreated CAUTIs in teaching hospitals compared to community sites (23.7% vs 10.8%; *P*=0.023) and in intensive care units (ICUs) than non-ICUs (27.1% vs 15.3%; *P*=0.012). An alternative diagnosis was documented in 59 of 63 untreated CAUTIs (93.7%), most often respiratory infection. In multivariable analysis, absence of pyuria was the strongest predictor of non-treatment status (adjusted odds ratio [aOR], 4.19; 95% CI, 1.92–9.14).

**Conclusions:** Most CAUTI events met the NHSN definition based on fever alone, and one in five was not treated, usually due to an alternative diagnosis. Alternative surveillance definition criteria, including lack of pyuria and antimicrobial treatment of UTI, warrant further evaluation.

## Introduction

Catheter-associated urinary tract infections (CAUTI) are a common preventable healthcare-associated infection^1^. For surveillance and public reporting, hospitals use the Centers for Disease Control and Prevention’s (CDC) National Healthcare Safety Network (NHSN) definition to identify cases^2^. The NHSN surveillance definition relies on a positive urine culture and the presence of a sign or symptom of infection, including fever. Fever causes some urine culture results to meet the CAUTI surveillance definition even when another source of fever is evident. This contrasts with clinical guidelines in which CAUTI is diagnosed only after other sources of infection are excluded.^3^

NHSN revised the CAUTI definition several times. The last revision in 2015 removed urinalysis criteria and raised the urine colony-count threshold to improve validity. These changes were associated with substantial reductions in reported CAUTI rates.^4^ However, concerns persist that the definition identifies a substantial number of cases of catheter-associated asymptomatic bacteriuria (CA-ASB) rather than infection.^5^

To address current gaps, we evaluated CAUTI events across a health system’s teaching and community hospitals, characterized opportunities for antimicrobial stewardship, and generated preliminary data that may inform future efforts to improve the validity of the CAUTI metric while rewarding judicious antibiotic use.

## Methods

### Study design and setting

We conducted a retrospective observational study of all CAUTI events reported to NHSN among adults (≥18 years) admitted to the Johns Hopkins Health System (JHHS) from January 1, 2023, through September 30, 2026. The study included five JHHS adult acute-care hospitals in Maryland and Washington, DC, region: two teaching hospitals (1,056 and 474 beds) and three community hospitals (253, 262 and 238 beds) (hospital characteristics are summarized in **Supplementary Table 1)**. Urinalysis with reflex to culture is available at the three community sites, while standalone urinalysis and urine cultures are available at all sites. Each hospital has its own antimicrobial stewardship and infection prevention program. A centralized infection prevention team performs CAUTI surveillance using current NHSN criteria. The Johns Hopkins Institutional Review Board deemed the study exempt (IRB00567676).

### Data collection and definitions

CAUTI events and the criteria that led them to meet the CAUTI surveillance definition were retrieved retrospectively from JHHS NHSN records. CAUTI criteria include a positive urine culture with ≥10^5^ colony-forming units/mL of no more than two species in a patient catheterized for more than two days, together with at least one qualifying sign or symptom (fever, suprapubic tenderness, or costovertebral angle pain or tenderness, or, after the catheter has been removed, urinary urgency, frequency, or dysuria). Two infection preventionists reviewed all cases to validate the qualifying criterion(a). Demographic characteristics, urinalysis and urine culture results were extracted from the electronic medical record. The indication for the urine culture, alternative sources of fever and the documented indication for antibiotics were determined by manual chart review. For each patient, multiple notes were reviewed, including on the date the culture was ordered and subsequent notes from both primary and consultant teams. When documentation of antibiotic indication was not found (“not documented”), the case was classified as “treated” to avoid underestimation of treatment for CAUTI. If a patient was started on an antibiotic that was subsequently stopped because the urine culture result was thought to represent asymptomatic bacteriuria, the case was counted as “untreated”.

A urinalysis was considered abnormal if any of the following was present: positive leukocyte esterase (trace or greater), positive nitrite, ≥10 white blood cells (WBC) per high-power field (HPF), or ≥4 red blood cells per HPF. Pyuria was defined as ≥10 WBC/HPF or >50 WBC/µL on automated urinalysis, or ≥10 WBC/HPF on manual microscopy when that was the only WBC result. CAUTIs were classified by NHSN location as ICU (critical care, specialty care area and step-down units) or ward (medical/surgical, specialty and oncology wards).

Immunocompromised was defined as absolute neutrophil count≥500 cells/mm^3^, HIV with CD4 <200, immunosuppressive therapy, or active chemotherapy.

### Statistical analysis

Continuous variables were compared with the Wilcoxon rank-sum test and categorical variables with the χ^2^ or Fisher exact test, as appropriate. Trends in the proportion of treated CAUTIs across the study period were assessed with the Cochran–Armitage test. We used multivariable logistic regression including age, sex, unit type (ICU vs non-ICU), hospital, immunocompromised status and pyuria status (pyuria, no pyuria, or no urinalysis or WBC result within 72 hours) to evaluate predictors of non-treatment. A two-sided P value < 0.05 was considered significant. Analyses were performed in Stata 16.1 (StataCorp, College Station, TX). We calculated a modified CAUTI standardized infection ratio (SIR) excluding treated patients from the numerator, and subsequently compared it to the NHSN CAUTI SIR.

## Results

### Cohort characteristics

We identified 301 CAUTI events, 236 (78.4%) at the two teaching hospitals and 65 (21.6%) at the three community hospitals. Characteristics of the cohort are summarized in **Table 1**. The median patient age was 68.7 years; 91 patients (30.2%) were immunocompromised, the median time from admission to CAUTI was 10 days (IQR, 5–24), and from indwelling urinary device insertion to infection was 8.5 days. Roughly half of the CAUTI events (47.8%) were attributed to ICUs/step-down units and 157 (52.2%) to wards. Nearly all events (95.7%) met a single sign or symptom criterion, and fever was the qualifying criterion in 244 (81.1%). Localized symptom findings were uncommon: suprapubic pain was recorded in 27 events (9.0%), dysuria in 24 (8.0%), urinary frequency in 11 (3.7%), and costovertebral angle pain in 9 (3.0%); no event qualified by suprapubic tenderness (**Suppl. Table 2**). A urinalysis was obtained within 72 hours before the event in 269 events (89.4%). Of these, 37 (13.75%) lacked pyuria. The median number of WBC/µL among those with pyuria was 835 (IQR 156, 2275) and 479 (43, 1532) for treated and untreated, respectively (**Table 1**)

**Figure.**
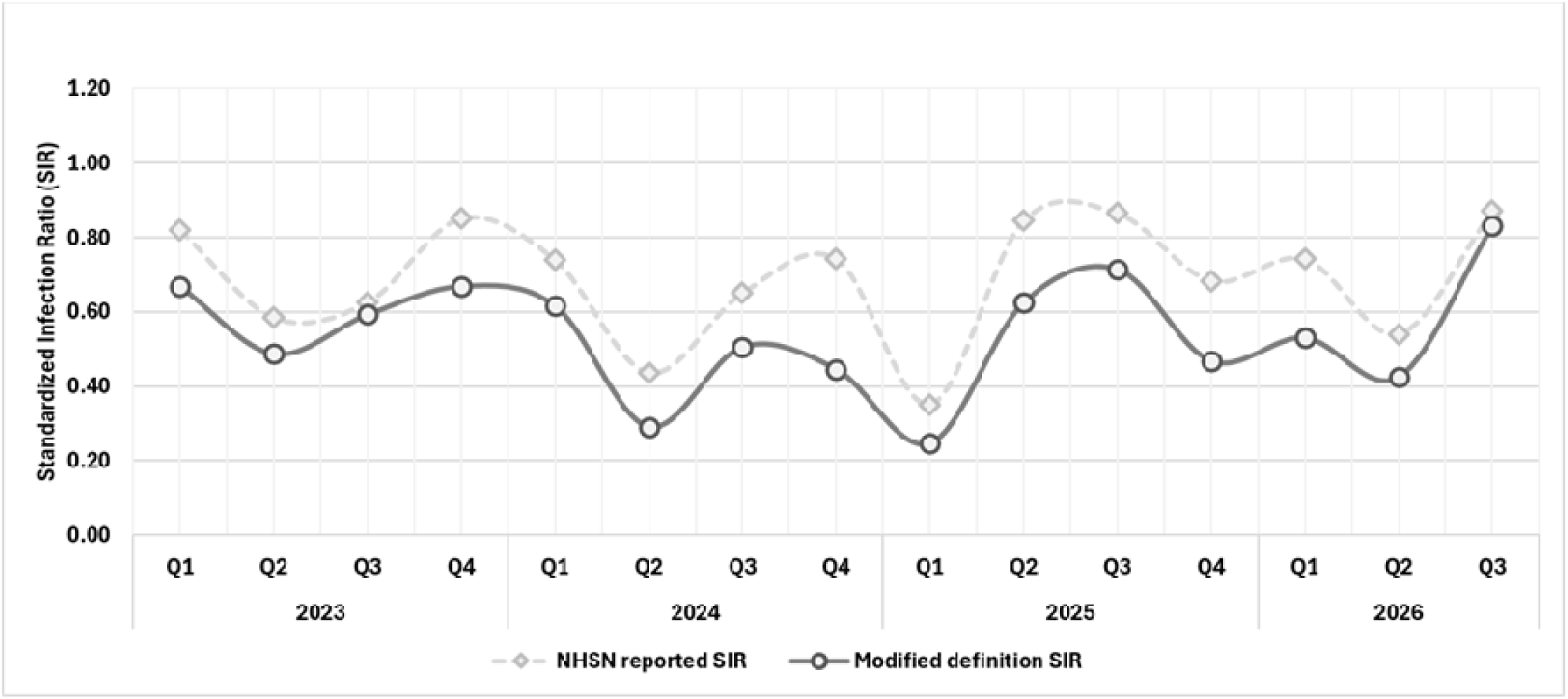
Adult catheter-associated urinary tract infection (CAUTI) standardized infection ratio (SIR) for participating hospitals. National Healthcare Safety Network reported CAUTI SIR (dotted line) and modified SIR after excluding treated CAUTI cases (solid line).

**Table 1.**
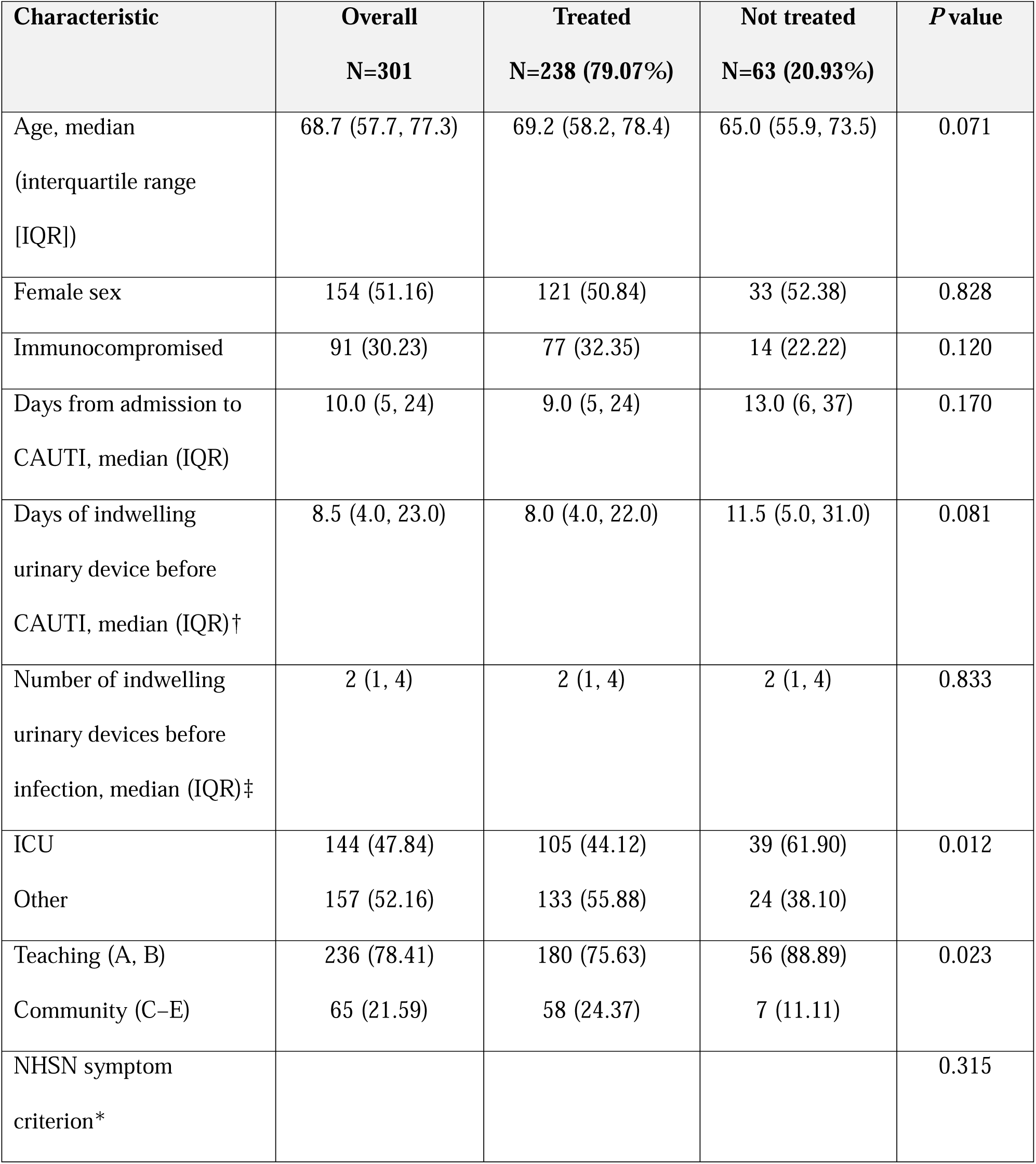
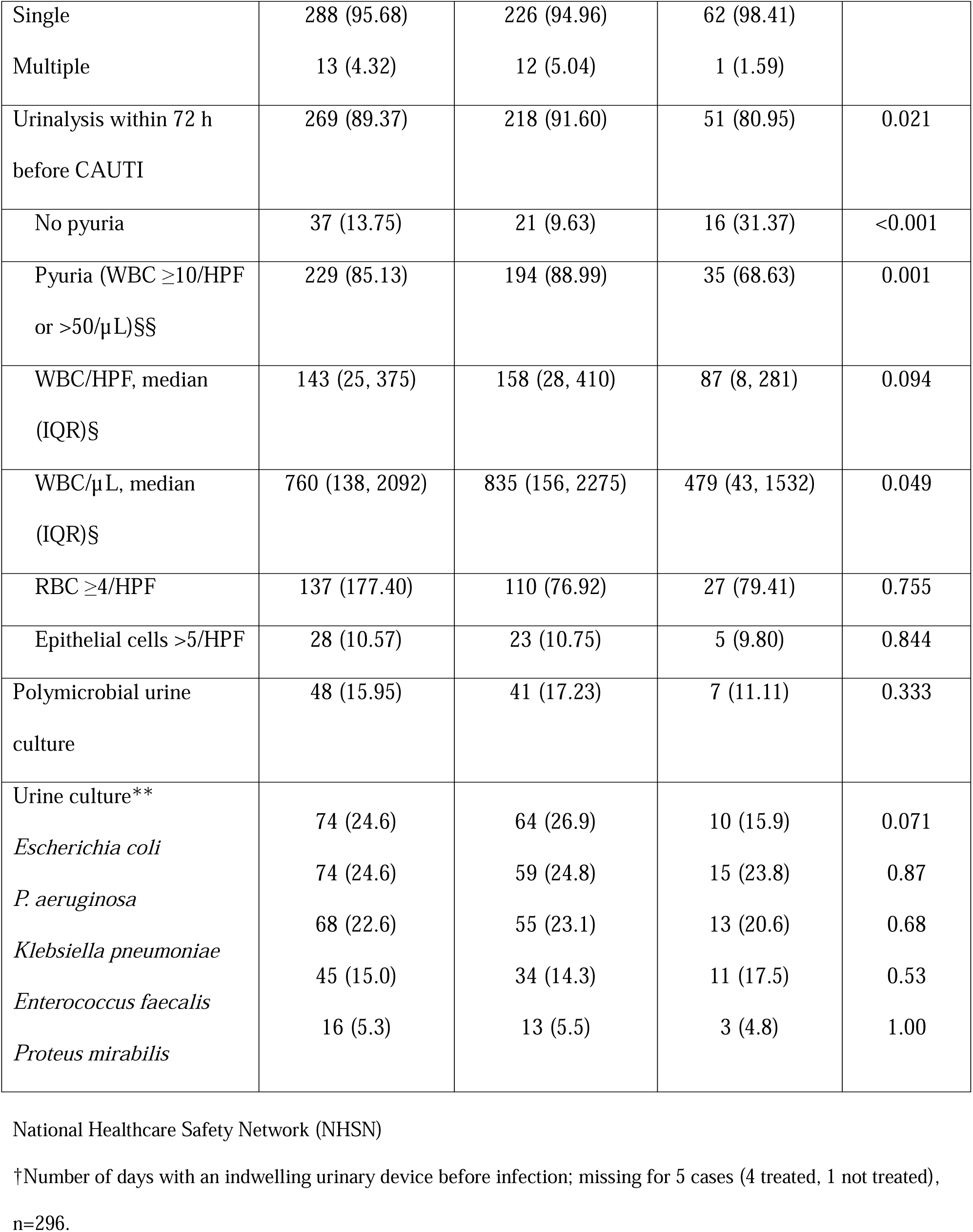
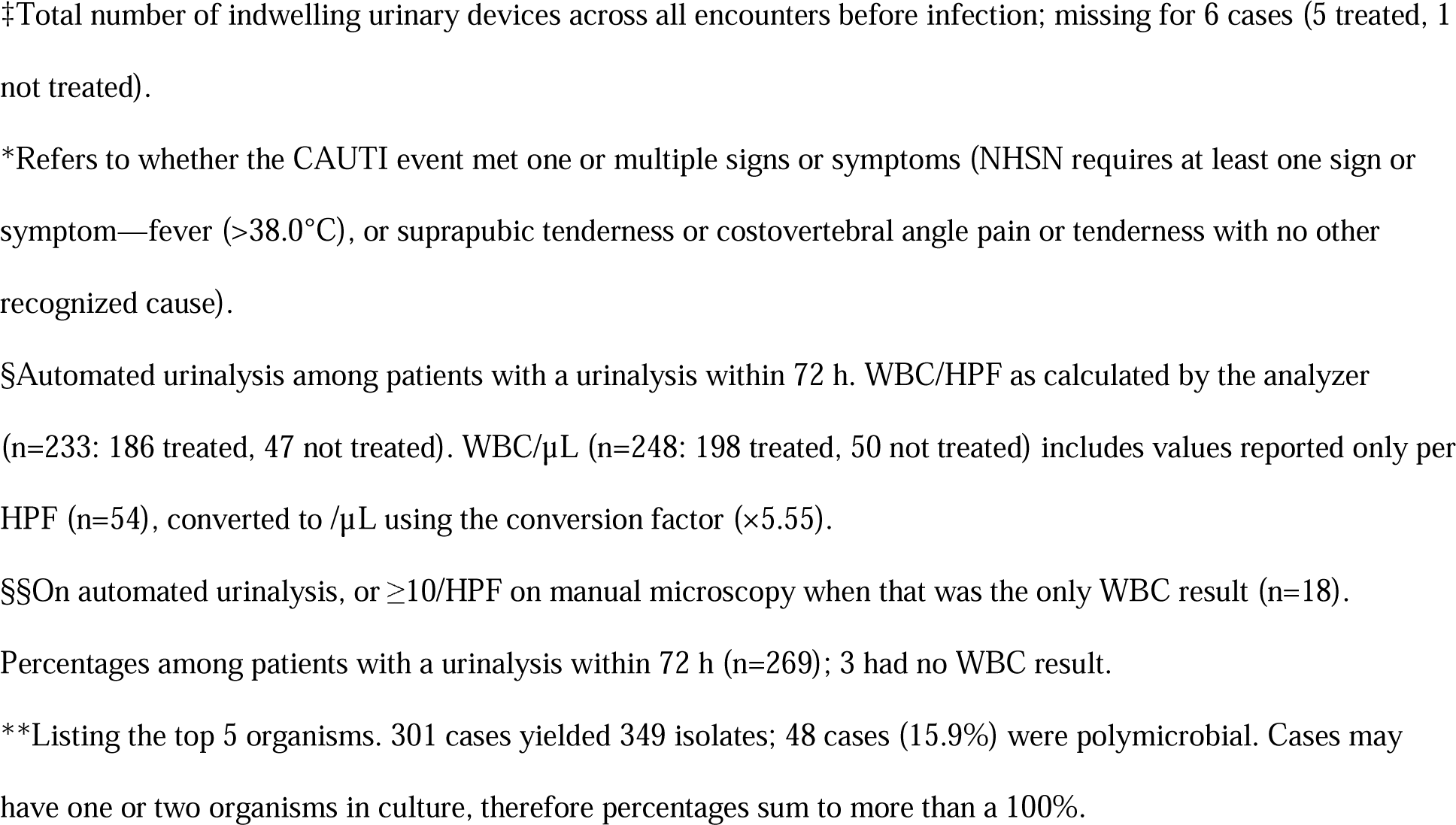
Characteristics of the cohort (adult patients admitted to JHHS, January 2023-May 2026, who met National Healthcare Safety Network [NHSN] Catheter-Associated Urinary Tract Infection [CAUTI] definition), overall and by treatment.

### Indications for urine culture and urine culture results

Indications for obtaining a urine culture documented by the treating team are shown in **Suppl. Table 3**. Fever was the most common indication (148, 49.2%), followed by leukocytosis (40, 13.3%), sepsis (22, 7.3%), and altered mental status (11, 3.7%). Urinary tract-specific indications were uncommon: dysuria in 22 events (7.3%), suprapubic, pelvic or abdominal pain in 17 (5.6%) and costovertebral angle pain or tenderness in 3 (1.0%). No indication was documented in 50 events (16.6%). Urine cultures yielded 349 isolates; 48 CAUTIs (15.9%) were polymicrobial. The most frequent organisms were *Escherichia coli* and *Pseudomonas aeruginosa* (74 each, 24.6%), *Klebsiella pneumoniae* (68, 22.6%), *Enterococcus faecalis* (45, 15.0%) and *Proteus mirabilis* (16, 5.3%).

### Antibiotic treatment and alternative diagnoses

Patient characteristics were similar among those who received antibiotics for CAUTI (n=238, 79/1%) and those who did not (n=63, 20.93%) (**Table 1**). Dysuria was more frequent among treated patients (9.7% vs 1.6%; *P*=0.036; **Suppl. Table 2**). The proportion of untreated CAUTIs varied by hospital, from 8.3% to 33.0% (**Suppl. Table 4**). Untreated patients were more frequent in ICUs than non-ICUs (27.1% vs 15.3%; *P*=0.012; **Suppl. Table 5**). Overall, the proportion of treated patients with CAUTIs did not change significantly over the study period (**Suppl. Figure 1**), but there was a significant trend towards lower treatment at hospital A, (−1.6 percentage points per quarter; 95% CI, −2.9 to −0.3; *P* =0.019) (**Suppl. Table 6**).

Untreated cases were approximately three times as likely to lack pyuria as treated ones (31.4% vs 9.6%; *P* <0.001). Of the 37 cases without pyuria, 16 (43.2%) were not treated, compared with 35 of 229 (15.3%) with pyuria. Among untreated cases, an alternative diagnosis was documented in primary or consultant team notes in 59 of 63 episodes (93.7%), most commonly respiratory infection (19, 30.2%), followed by central fever (9, 14.3%), burn injury (7, 11.1%), skin, soft tissue or bone infection (6, 9.5%) and other noninfectious causes (6, 9.5%) (**Suppl. Table 7**).

In multivariable logistic regression, after adjusting for age, sex, unit type, hospital and immunocompromised status, absence of pyuria was the strongest independent predictor of non-treatment (aOR, 4.19; 95% CI, 1.92–9.14). Non-treatment was more likely at the second teaching hospital (B; aOR, 2.05; 95% CI, 1.03–4.07) than at the reference teaching hospital (A), and in patients without a urinalysis or WBC result within 72 hours (aOR, 2.61; 95% CI, 1.10–6.18). (**Table 2**).

**Table 2.** Multivariable logistic regression model of predictors of not being treated for a National Healthcare Safety Network (NHSN)-reported catheter-associated urinary tract infection (CAUTI), N=301.

| **Variable** | **Unadjusted OR (95% CI)** | ***P value*** | **Adjusted OR (95% CI)** | ***P value*** |
|---|---|---|---|---|
| Pyuria status |  |  |  |  |
| No pyuria | 4.22 (2.01–8.88) | <0.001 | 4.19 (1.92–9.14) | <0.001 |
| No UA within 72 h of CAUTI | 2.89 (1.32–6.34) | 0.008 | 2.61 (1.10–6.18) | 0.029 |
| Age, per 10 years | 0.87 (0.74–1.02) | 0.094 | 0.92 (0.77–1.10) | 0.352 |
| Female | 1.06 (0.61–1.85) | 0.828 | 1.23 (0.65–2.34) | 0.524 |
| Immunocompromised | 0.60 (0.31–1.15) | 0.122 | 0.81 (0.39–1.69) | 0.579 |
| ICU | 2.06 (1.16–3.64) | 0.013 | 1.70 (0.90–3.20) | 0.100 |
| Hospital |  |  |  |  |
| B | 2.20 (1.20–4.05) | 0.011 | 2.05 (1.03–4.07) | 0.041 |
| C | 0.41 (0.12–1.43) | 0.160 | 0.41 (0.11–1.50) | 0.177 |
| D | 0.90 (0.24–3.31) | 0.870 | 0.98 (0.24–3.97) | 0.981 |
| E | 0.45 (0.06–3.65) | 0.453 | 0.39 (0.04–3.42) | 0.392 |

OR: odds ratio. UA: urinalysis. Reference categories: pyuria, male sex, non-ICU and hospital A. ICU includes critical care, specialty care and step-down units. Pyuria as defined in Methods.

### Potential impact of treatment decisions on CAUTI rates

To evaluate the potential impact of excluding untreated patients (i.e., those deemed to have CA-ASB) from the CAUTI surveillance definition, we recalculated facility-wide CAUTI modified SIR excluding these cases. For each year, the reduction from reported SIR to modified SIR was: 0.72 to 0.61 (2023), 0.65 to 0.47 (2024), 0.68 to 0.51 (2025), and 0.72 to 0.59 (2026) (**Figure and Suppl. Figure 2 and 3).**

## Discussion

In this multicenter cohort, one in five CAUTIs reported to NHSN were not treated with antibiotics. This indicates that the clinical teams interpreted the positive urine culture as representing CA-ASB, yet each event counted towards the hospital’s reported CAUTI rate and standardized infection ratio. Most CAUTI events were diagnosed based on fever and a positive culture alone, and absence of pyuria was the strongest predictor of withholding antibiotics. These findings illustrate two limitations of the current NHSN CAUTI definition; it counts events that clinicians do not consider infections, and it does not reward appropriate diagnosis and non-treatment of CA-ASB.

Our findings are consistent with prior reports of poor agreement between CAUTI surveillance criteria and clinical diagnosis. An earlier version of the NHSN CAUTI definition had a positive predictive value (PPV) of only 35% when compared with the assessment of infectious diseases consultants.^6^ In a recent single-center review of 143 NHSN-reported CAUTIs, 40% of cases were deemed CA-ASB (70% had a confirmed alternative source of fever or leukocytosis, most commonly lower respiratory tract infection) and 18% met prespecified criteria for clinical CAUTI which the authors defined as bacteremia with a matching urinary organism, fever or leukocytosis with urinary signs or symptoms, urinary or pelvic symptoms, or supportive imaging; ^5^ In that study, most patients received antibiotics. The larger proportion of untreated events in our cohort may reflect local antimicrobial stewardship efforts or differences in patient populations.

Prior studies evaluating the NHSN CAUTI metric have focused largely on the denominator and on surveillance automation. Fakih and colleagues used computer-based Monte Carlo simulation to compare CAUTI rates using the NHSN-recommended catheter-days with patient-days. The authors found that the population-based rates better captured the benefit of catheter avoidance than rates per catheter-day.^7–9^ Electronic algorithms reproduce the NHSN definition reliably and efficiently.^10–14^ Evaluations of the numerator have been limited to official revisions of the definition^4,15^ and single-center projections.^16^ An ideal CAUTI measure would more closely reflect clinical definitions of urinary tract infections requiring treatment, while excluding positive urine culture results that are not treated because they are deemed to be CA-ASB. It would also utilize patient-days in the denominator. These changes would focus the surveillance definition on clinically significant infections while excluding cases that appropriately minimized or avoided unnecessary use of urinary catheters and antibiotics.

Pyuria is common in catheterized patients due to urothelial irritation and bacterial colonization. The negative predictive value of lack of pyuria (<5 WBC/HPF) is high (93%) for the absence of clinician-diagnosed UTI among men with indwelling catheters.^17^ Simpson et al. reported 19% of CAUTIs could have been avoided by resetting a pyuria cut-off of 10WBC/HPF from 5 WBC/HPF.^16^ In our cohort, 13.8% of events with a urinalysis lacked pyuria, and these events had about four times the odds of being untreated.

Fever is common and nonspecific in hospitalized patients, occurring in up to 20% of hospitalized adults (or up to 80% in those in the intensive care); yet approximately half of these fevers are not indicative of an infectious process.^18,19^ Consistent with this, our study found that untreated events were more common in ICUs, and neurologic causes and burn injury were among the most frequent alternative diagnoses documented by clinicians. Including antimicrobial treatment as part of the CAUTI surveillance definition could address this issue. NHSN has proposed a healthcare facility-onset, antibiotic-treated *C. difficile* infection (HT-HO CDI) definition, under which a laboratory-identified positive specimen counts only if accompanied by at least 5 consecutive days of anti-CDI therapy.^26,27^ A similar treatment-based CAUTI definition would have excluded 21% of events in our cohort. Unlike CDI, antibiotics commonly used to treat UTI are often given for other infections;^5^ therefore, the indication for antibiotics in treated patients may need to be assessed. In this study, no clinical indication was documented for most urine cultures, highlighting a need for improved diagnostic stewardship. Diagnostic stewardship efforts for urine cultures have focused on reducing inappropriate cultures as part of “pan-culture” for fever in the ICU;^21^.^24^ In the future, large language models may help identify patients who may benefit from urine cultures.

This study has limitations. It was retrospective, and antibiotic indications relied on clinical documentation, which may be incomplete and may over- or underestimate clinical urinary tract infections. Review of notes from multiple teams on multiple days helped mitigate this limitation. Cases with undocumented treatment status were classified as treated to avoid overestimation of untreated cases. Finally, we did not account for differences in urine culturing practices, case mix, or antimicrobial or diagnostic stewardship practices across hospitals, all of which should be considered in future work to improve the CAUTI surveillance definition.

In this study, most CAUTI events met the NHSN definition based on fever alone, and one in five was not treated, usually due to an alternative diagnosis. Findings from this study could inform future work to optimize the NHSN CAUTI metric. Alternative surveillance definition criteria, including lack of pyuria, and antimicrobial treatment of UTI warrant further evaluation. A metric that better reflects clinical infection and antimicrobial stewardship is needed.

## Acknowledgments

The authors would like to thank James Lillie for his valuable contribution to data extraction.

## Financial support

VF, SEC and SCK received funding from the Prevention Epicenters Program of the Centers for Disease Control Prevention (grant number 6 U54CK000617-05-03). The study contents are solely the responsibility of the authors and do not necessarily represent the official views of CDC.”

## Competing interests

All authors report no conflicts of interest relevant to this article.

## Data availability

The data that support the findings of this study are available from the corresponding author upon reasonable request.

## Notes

### Competing Interest Statement

The authors have declared no competing interest.

### Author Declarations

The Johns Hopkins Institutional Review Board deemed the study exempt (IRB00567676).

